# Feasibility and Acceptability of a Prospective Remote Pilot Study for Assessment of Airborne Environmental Exposures and Male Fertility

**DOI:** 10.64898/2026.11.03.26367192

**Authors:** Ingrid H. Robertson, Liddy Boland, Damon Wing Hey Chan, Maximilian S. Jentzsch, Kathleen Moloney, Amanda J. Haack, Madeleine Eakman, Filip Stefanovic, Lauren G. Brown, Charles Tilander, Christine Loftus, Jane Edelson, Charles H. Muller, Erwin Berthier, Krystal J. Godri Pollitt, Nicole A. Errett, James F. Smith, Catherine J. Karr, John Amory, Ashleigh B. Theberge, Tristan Nicholson

**Author notes:** IR and LB contributed equally to this work.

## Abstract

**Objective:** To evaluate the feasibility and acceptability of serial biosampling and environmental exposure assessment in reproductive-aged men and compare participant experiences with in-person versus remote sample collection.

**Design:** Prospective cohort pilot study.

**Subjects:** Eighteen healthy men aged 21–44 years with no history of infertility, medical conditions or medications known to cause infertility, or prior scrotal or testicular surgery.

**Exposure:** Indoor and personal environmental exposure monitoring using PurpleAir monitors and Fresh Air wristbands, respectively.

**Main Outcome Measures:** Feasibility of repeated remote semen and blood collection, evaluated by participant retention and number of samples returned, and participant-reported acceptability, ease of use, discomfort, preference for at-home versus in-clinic sampling, and willingness to participate in longitudinal studies.

**Results:** 18 participants completed the initial in-person and scheduled at-home sampling, and 16 (89%) completed subsequent remote sampling. Participants successfully collected and shipped serial capillary blood samples using homeRNA and semen samples using the Fellow Semen Analysis Kit. Participants rated both collection methods as easy to use and reported minimal discomfort with blood collection. At-home participation was preferred over in-clinic sampling, particularly for semen collection. All 16 participants completing the final survey reported willingness to participate in a similar study again. Participants were willing to provide a median of 18 semen samples (range, 2–40) and 18 blood samples (range, 5–40) over a future hypothetical study. Both home-based environmental monitoring approaches were well accepted.

**Conclusion:** Serial remote semen and blood collection paired with environmental exposure monitoring was feasible and acceptable among reproductive-aged men. High retention, preference for at-home sampling, and willingness to provide repeated samples support this approach for larger longitudinal studies investigating environmental exposures and male reproductive health.

## INTRODUCTION

Environmental influences, such as air pollution, are increasingly recognized as a major contributor to male reproductive health. Air pollution exposure has been associated with adverse effects on sperm morphology,(1–4) motility,(1,2,5) count,(6,7) and concentration,(6,8,9) as well as semen volume.(5,6) Indoor sources of air pollutants can result from daily activities, including cooking, cleaning, or smoking; whereas outdoor exposures originate from anthropogenic sources including combustion of fossil fuels, agricultural and construction activities, as well as other sources such as wildfires.(10–12) Wildfire smoke exposure has in particular increased in recent years with many regions experiencing episodic exposures.(13,14) While commonly assessed as particulate matter of 2.5 microns or smaller (PM_2.5_), wildfire smoke also includes compound groups such as polycyclic aromatic hydrocarbons (PAHs), which have been associated with an increase in azoospermia, the absence of sperm cells in semen, and a reduction in semen volume.(5) Further, exposure to wildfire smoke is associated with decreased sperm concentration, decreased total motile sperm count, decreased percent normal heads, and increased odds of azoospermia.(15–17) Given the complex composition of wildfire smoke and co-exposures with indoor air pollution, studies that capture the dynamics of human exposures and follow individuals over time are important to understand the impact of airborne exposures on human sperm quality.(18) Here, we present a feasibility and acceptability pilot study evaluating participant perception and willingness to perform at-home dual biospecimen collection (semen and blood) paired with at-home/wearable exposure devices to investigate the effects of wildfire smoke and daily environmental exposures on semen quality and systemic response.

Recent advancements in at-home biofluid collection, including blood(19–22) and semen sampling,(23–25) and wearable device(26,27) technology have expanded research possibilities beyond the clinic. In this study we used the homeRNAmax Blood Collection and Stabilization Kit (referred to throughout as homeRNA),(28) a self-collection platform for peripheral blood and RNA stabilization, and the Fellow Semen Analysis Kit (referred to throughout as Fellow), a mail-in semen analysis platform.(25) A previous validation study compared semen samples from the same participant that were analyzed within one-hour to mailed, and they were found to be highly correlated in concentration, motility, and morphology.(25) Similarly, several previous validation studies of the homeRNA platform show excellent cellular RNA stabilization across a range of temperatures, including summer months and during wildfire smoke events.(29–32) Being able to measure peripheral blood transcriptomic signatures in addition to sperm quality would enable research on the systemic effects of exposure (e.g., inflammation) that may underlie effects on fertility.

Considering the importance of studying the impact of airborne environmental exposures on male fertility, we aimed to [1] develop a remote, adaptable sampling protocol that captures ubiquitous environmental exposures paired with biospecimen measurements specifically designed for rapid deployment during short-lived exposure events such as wildfire smoke, [2] ascertain the feasibility of multimodal, longitudinal environmental exposure assessment and biospecimen collection (e.g., retention, number of samples returned), and [3] evaluate acceptability of the remote protocol through participant surveys. To do this, we enrolled reproductive age men in a dual clinical/remote study in which participants self-collected blood using the homeRNA platform and semen using the Fellow kit, continuously monitored home indoor PM_2.5_ mass concentration estimates using PurpleAir monitors, and, in a subset of participants, passively sampled environmental organic chemical exposures continuously over a seven day period using Fresh Air wristbands. By combining these remote collection protocols, we evaluated the feasibility of a multi-biofluid collection study with multiple timepoints where, during wildfire season, enrolled participants provided one baseline sample in-clinic and up to three samples remotely. Our study leverages new approaches that enable participant biosample collection and personal exposure sampling to create a framework for future large studies aimed at advancing scientific questions on wildfire smoke exposure and male fertility.

## MATERIALS AND METHODS

### PERSONAL BIOSPECIMEN COLLECTION AND EXPOSURE MONITORS

The *Fellow Semen Analysis Kit* (Fellow) is a semen analysis platform that enables at-home semen collection, stabilization, and shipment for laboratory semen analysis (SI.1).(24,25) The *homeRNAmax Blood Collection and Stabilization Kit* (homeRNA) combines a commercially available peripheral blood collection device with a custom-engineered stabilization tube (SI.2).(28–34) *Fresh Air* wristbands (FAW) are wearable passive samplers that provide time-integrated measurements of personal exposure to a range of airborne environmental contaminants (SI.3).(35–37) Participants were also provided with *PurpleAir Touch* monitors to measure indoor PM_2.5_ mass concentration(SI.4).(38–41)

### STUDY POPULATION AND DESIGN

#### Enrollment

This study was approved by the University of Washington Institutional Review Board (IRB; protocol 00022559). Participants were recruited through social media advertisements and completed an eligibility survey REDCap). Eligibility criteria included being 18–65 years of age, having no prior infertility diagnosis, being able to produce a semen sample through ejaculation, and willingness to use an air-monitoring device. Of the 36 individuals who completed the survey, 35 met the eligibility criteria. Of these, twenty eligible individuals were randomly selected. All participants provided informed consent.

#### Enrollment Kits

Participants were shipped study kits via UPS Next Day Air containing (1) an indoor PurpleAir monitor, a setup guide, and (3) a pre-labeled UPS LabPak for sample return. Upon receiving the kit, participants were instructed to install the PurpleAir monitor in their home (any room except kitchen) and operate it continuously for the duration of the study. Half of the enrolled participants (n=9) were randomly selected to also receive a FAW for personal airborne exposure monitoring. Participants who received a FAW were instructed to store it in the freezer, then remove and wear it continuously beginning 7 days before their initial study visit.

#### At-clinic pre-event

All participants attended one in-person visit at the University of Washington Seattle campus (Figure 1A). Participants were instructed to abstain from ejaculation for 2-5 days prior to the in-person visit. For participants who wore a FAW, study staff collected the wristband, transferred the sample case to a glass jar, and shipped it by FedEx to the Pollitt Laboratory at Yale University for analysis.

**Figure 1.**
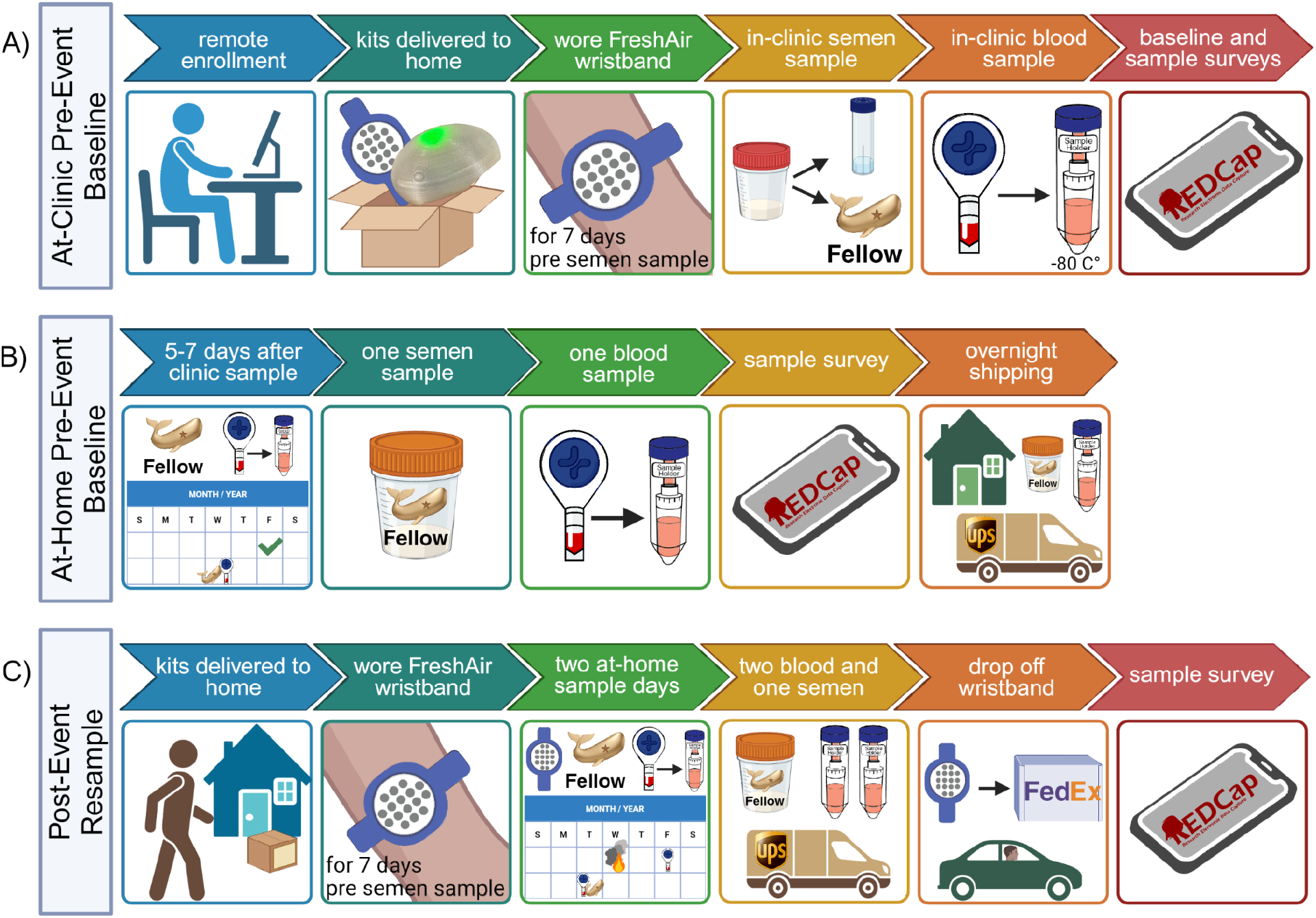
Study design and sampling workflow. **(A)** The At-Clinic Pre-Event Baseline included remote enrollment, delivery of study materials, in-clinic semen and homeRNA blood collection, and surveys. Half of participants were randomly selected to wear Fresh Air Wristbands. **(B)** The At-Home Pre-Event Baseline consisted of a homeRNA and Fellow kit 5-7 days after the clinic visit. **(C)** The Post-Event Resample consisted of two at-home sampling days, including two homeRNA blood samples and one Fellow semen sample, followed by surveys. Participants assigned a Fresh Air wristband wore it for seven days before sampling and returned it after collection. Created in https://BioRender.com

At the visit, participants produced a semen sample without lubricant, and a semen analysis was performed according to *WHO* guidelines.(42) An aliquot of well-mixed semen of known volume was transferred to a Fellow specimen container by study staff, combined with the provided preservation solution, and shipped to the Fellow lab in San Francisco for analysis.

After watching an instructional video, participants self-collected a blood sample using the homeRNA kit. They then completed a survey assessing environmental exposures, smoking habits, perceived stress, comfort with sampling during a disaster, and their experiences with the Fellow and homeRNA kits. Experiences were rated using 5-point Likert scales, and participants reported any challenges encountered. Before leaving, participants received a take-home kit containing the materials needed to repeat the semen and blood collections 5–7 days later. The sampling date was selected in coordination with study staff.

#### At-home pre-event

Five to seven days after the in-person visit (and after abstaining from ejaculation for 2-5 days), participants collected semen and blood samples at home using the Fellow and homeRNA kits. Participants received reminders by text message and email 24 hours before and on the scheduled sampling date. A UPS courier collected both samples on the day of collection. Fellow kits were shipped to Fellow for analysis, and homeRNA kits were shipped to the University of Washington (Figure 1B).

Participants completed a post-sampling survey assessing ease of use, comfort with each collection platform and setting, recent exposure to poor outdoor air quality, smoking habits, and recreational activities. They were informed that additional sampling could be initiated in response to a wildfire smoke event and were instructed to keep their PurpleAir monitors operating throughout the study.

#### Post-event resampling

The goal of the timepoint was to capture biospecimens and exposure during and after a wildfire smoke event. The study team monitored local air quality data and forecasts. A qualifying wildfire smoke event was defined as a forecasted Air Quality Index of moderate or higher for at least two consecutive days, based on the U.S. Environmental Protection Agency AirNow website. One participant met the predetermined threshold for a wildfire event (Bear Gulch, July 2025)(43) and resampling proceeded as planned. While a low level of smoke from the fire was experienced by the rest of the participants, it did not meet the study’s predetermined threshold, nor did another event prior to the conclusion of wildfire smoke season in the region. Therefore, the study team initiated resampling steps to evaluate the feasibility of repeated sampling rather than in response to a qualifying event.

Before repeat sampling, participants previously assigned a FAW received a second wristband and were instructed to store it in the freezer until notified. Participants received kits containing the materials required for two homeRNA collections and one Fellow collection (Figure 1C). Participants assigned a FAW were instructed to begin wearing it when repeat sampling was initiated. Five days after initiation, participants collected the first homeRNA sample and shipped it to the study team on the same day. Two to 3 days after, participants collected using homeRNA and Fellow, shipping overnight to respective labs.

## RESULTS

### High participant retention across multiple sampling timepoints

Participant enrollment was completed in 2 days (Table1). Two of the 20 enrolled participants withdrew before completing the initial study activities. The remaining 18 participants completed PurpleAir monitor setup, surveys, in-person pre-event baseline, and at-home pre-event semen and blood baseline collection 5–7 days later. Nine participants also wore a FAW. Sixteen participants completed sampling and surveys during the post-event resample. Of the nine participants assigned to wear FAWs, six returned their post-event wristband.

**Table 1.** Participant demographic characteristics, pre-existing conditions, and seasonal allergies. Data are presented as n(%), N=18.

| DEMOGRAPHICS |  | ALLERGIES |  |
|---|---|---|---|
| **Age** | **N=18(%)** | **Do you have seasonal allergies or hay fever?** | **N=18(%)** |
| 20-29 | 6(33) | Yes | 10(56) |
| 30-39 | 8(44) | No | 8(44) |
| 40-49 | 4(22) | **Is there a time of year in which your allergies are worse?** |  |
| 50-59 | 0 | Spring | 7(39) |
| 60-69 | 0 | Fall | 1(6) |
| **Race** |  | Summer | 3(17) |
| American Indian or Alaskan Native | 1(6) | Winter | 0 |
| Asian | 8(44) | **What are your typical seasonal allergy symptoms?** |  |
| Black or African-American | 0 | Runny Nose | 6(33) |
| Native Hawaiian or Other Pacific Islander | 0 | Stuffy Nose | 6(33) |
| White | 8(44) | Itchy Mouth or Throat | 2(11) |
| Other | 2(11) | Sneezing | 6(33) |
| **Are you Hispanic, Latino, or (of) Spanish origin?** |  | Wheezing | 1(6) |
| Yes | 3(17) | Shortness of Breath | 1(6) |
| No | 15(83) | Cough | 1(6) |
| **Area** |  | Rashes | 2(11) |
| Urban | 18(100 | Fatigue | 1(6) |
| Rural | 0 | Headache | 0 |
|  |  | Nausea or Vomiting | 0 |
| PRE-EXISTING CONDITIONS |  | Fever | 0 |
| **Has your doctor or medical provider ever told you that you have any of the following conditions?** | **N=18(%)** |  |  |
| Cystic Fibrosis | 0 |  |  |
| Asthma | 4(22) |  |  |
| Lung Disease | 0 |  |  |
| Hypertension/High Blood Pressure | 2(11) |  |  |
| Hyperlipidemia/Hypercholesterolemia | 2(11) |  |  |
| Cholesterol | 3(17) |  |  |
| Other Heart Disease | 0 |  |  |
| Anemia | 0 |  |  |
| Liver Disease | 0 |  |  |
| Diabetes | 0 |  |  |
| Cancer | 0 |  |  |
| Chronic Kidney Disease | 0 |  |  |
| Other Chronic Condition | 0 |  |  |
| None | 12(67) |  |  |

### Positive perceptions and preference for remote semen collection

Participants consistently reported that the semen collection procedure was easy to perform, the written instructions were easy to understand, and they would be willing to perform this type of collection again (Figure 2A,B). Fifteen of 16 participants preferred remote semen collection. In free-response questions, participants cited greater privacy, convenience and flexibility with remote collection, avoiding travel and the potential discomfort with collection in a clinical setting.

**Figure 2.**
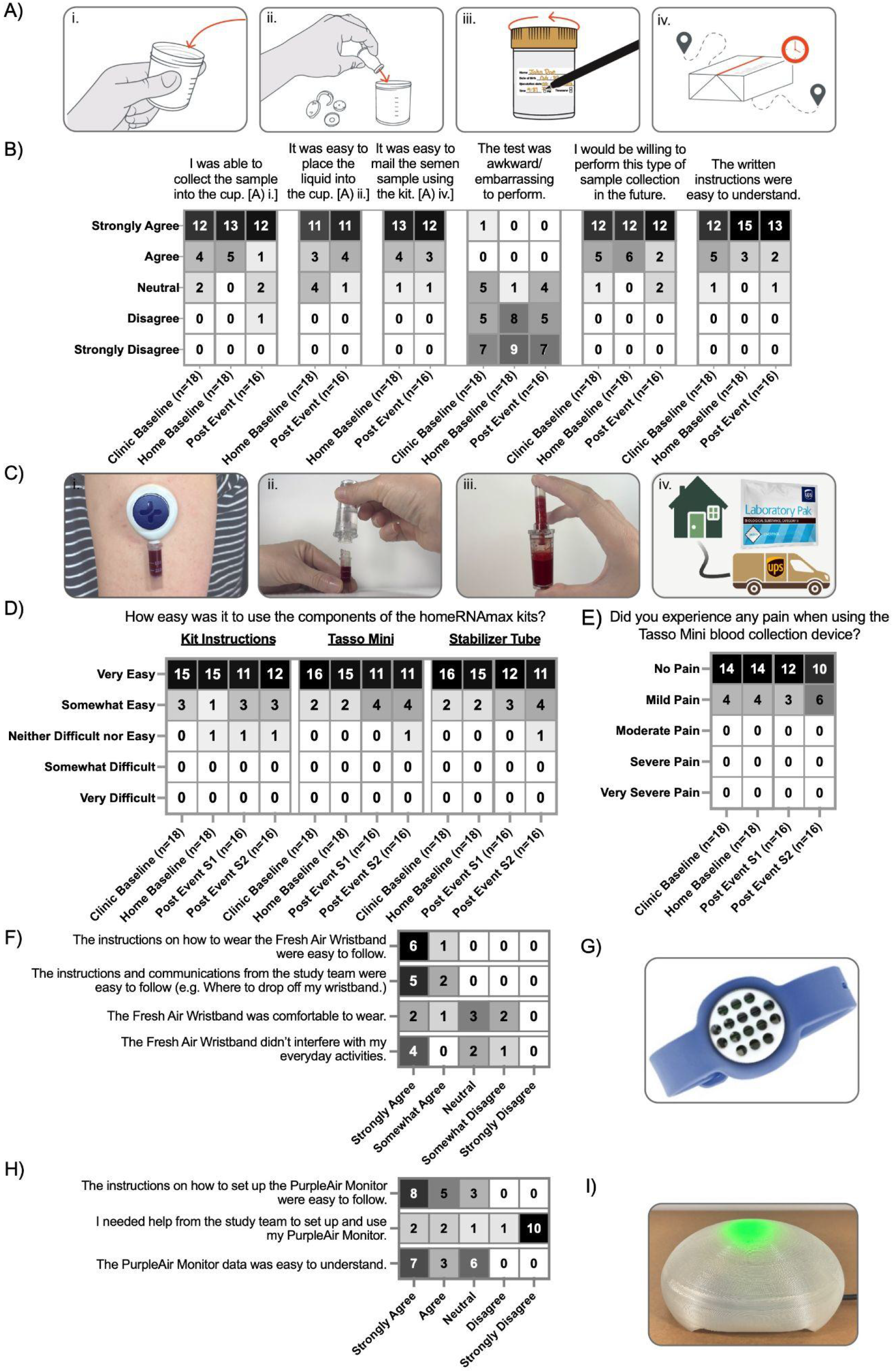
Participant experience with sampling kits and environmental exposure monitoring devices. **(A i–iv)** Fellow Semen Analysis Kit workflow: (i) semen collection, (ii) stabilization, (iii) sealing and labeling, and (iv) overnight shipment. **(B)** Participant-reported semen collection experience at At-Clinic Pre-Event Baseline (Clinic Baseline) and At-Home Pre-Event Baseline (Home Baseline) and Post-Event Resample (Post Event). **(C i–iv)** homeRNAmax Blood Collection and Stabilization workflow: (i) Tasso-Mini blood collection, (ii) connection to the stabilization tube, (iii) mixing with stabilizer, and (iv) overnight shipment. **(D–E)** Participant-reported ease of use (D) and pain (E) with homeRNA blood collection. Two homeRNA samples were collected during the Post-Event Resample (Post-Event S1 and S2). **(F–G)** Participant-reported experience with (F) and image of (G) the Fresh Air Wristband. **(H–I)** Participant-reported experience with (H) and image of (I) the PurpleAir monitor; green indicates “Good” PM2.5 Air Quality Index (AQI).

### Participants considered remote blood collection easier than in-person collection

Sixteen out of 18 participants completed the closing survey. Most of these participants(14 of 16) indicated a preference for remote blood sampling due to convenience and all participants reported mild to no pain (Figure 2C-E). Participants reported that traveling to a clinic would disrupt their schedules, whereas at-home sampling allowed them to collect on their own time.

### Participants reported willingness to participate in multiyear longitudinal studies

All participants who completed the closing survey (n=16) reported that they would participate in a similar study again (Figure 3A-C). Among these 16 participants, 12 indicated willingness to participate in a remote semen collection study involving approximately six samples per year for more than five years. The remaining four were willing to participate for at least one year. For remote blood collection involving approximately 15 samples per year, 10 participants were willing to participate for more than five years, while the remaining six were willing to participate for at least one year. Participants were also asked how many samples they would be willing to collect during a nine-month study. For semen, responses ranged from 2 to 40 samples (median, 18; mean, 16). For blood, responses ranged from 5 to 40 samples (median, 18; mean, 17).

**Figure 3.**
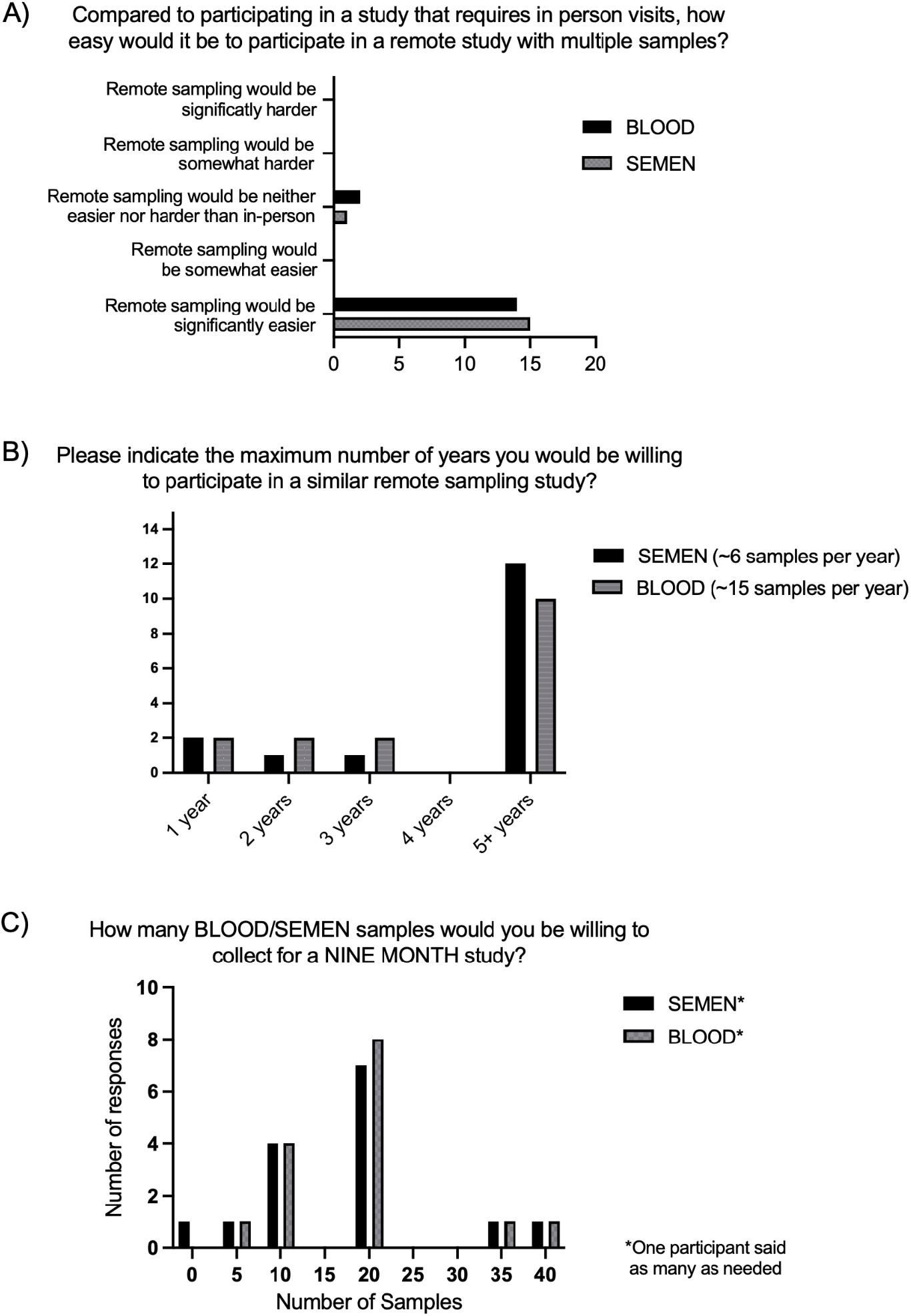
Participant preferences and willingness to participate in longitudinal studies involving repeated semen and blood collection, n=16 participants responding to closing survey. **(A)** Participant preference for at-home versus in-clinic study participation. **(B)** Willingness to participate in a longitudinal study requiring approximately six semen and 15 blood samples annually. **(C)** Number of semen and blood samples participants were willing to provide over nine months. Median willingness was 18 samples for both semen (range: 2–40) and blood (range: 5–40).

### PurpleAir monitors were easy to use but prompted few behavioral changes

All 18 participants were provided a PurpleAir monitor. From the closing survey, 6 of 16 participants used the PurpleAir mobile application or website to obtain information beyond the monitor’s color indicator, and two enabled poor-air-quality notifications or alerts. When asked what actions they took when the indicator changed from green to yellow or above, six participants reported closing their windows, while nine reported taking no mitigating actions. Fifteen participants indicated that they would participate in a similar study using a PurpleAir monitor. Thirteen were willing to participate in a study using two monitors—one indoors and one outdoors—while three indicated that they might participate but would require additional information (Figure 2H-I).

### Environmental exposures were captured across participants

Indoor PM_2.5_ mass concentration measurements were collected from 17 participants (94%), data from one participant was not available. Outdoor measurements were estimated for all 18 participants (100%) and were reported elsewhere. During the measurement period, smoke from the Bear Gulch Fire was detectable among enrolled study participants, coinciding with a small but significant increase in indoor and outdoor PM_2.5_ mass concentrations; however, for all but one participant, it did not meet our study’s prespecified sampling threshold for a wildfire smoke event. Participant experiences for the FAW are in Figure 2.F,G. For more details on exposure captured by the FAW please see Supplementary Information (SI.5-7).

### Study staff interactions with participants

Across the study (N=18), participants received an average of 2.95 phone calls (SD, 3.39) and 12.8 text messages (SD, 12.82) from study staff. Staff noted greater difficulty corresponding with participants affiliated with educational institutions during examination periods and the beginning of the academic term.

## DISCUSSION

This pilot study establishes the feasibility of a larger future study remotely examining the acute effects of environmental factors on male fertility. The aims were to develop a remote multi-biofluid collection protocol paired with environmental exposure measurements that can be deployed quickly during a trigger event, to determine the feasibility of a longitudinal multi-specimen study, and to evaluate the acceptability of remote participation. This was achieved through combining at-home mail-in blood and semen collection with passive monitoring of PM_2.5_ and airborne organic chemical exposures for individual participants. Participants reported both biosampling modalities (Fellow and homeRNA) were easy to use, more comfortable and preferable at-home versus in-clinic, and appealing options for collecting multiple samples over the course of a multiyear study.

Many studies of male reproductive health recruit participants in the context of pregnancy planning or fertility evaluation. For example, in the Pregnancy Study Online (PRESTO), female participants enroll first and subsequently invite their male partners to participate, with a subset of male partners completing two at-home semen tests.(44,45) Other studies, including those by Radwan et al., have recruited men already attending fertility clinics for diagnostic evaluation.(4,15) In contrast, this study recruited reproductive-age men regardless of relationship status, fertility concerns, or reproductive intentions. This broader recruitment approach enabled the inclusion of men who may not otherwise participate in fertility-focused research. Expanding recruitment beyond couples planning a pregnancy and patients seeking fertility care may improve the generalizability of male reproductive health research and provide a more comprehensive understanding of semen quality in the broader population. This is particularly relevant because semen quality has been associated with overall health, future disease risk, mortality, and life expectancy, suggesting that male reproductive health may reflect health outcomes beyond fertility alone.(46,47)

Recruitment was completed within 72 hours of study launch, with high retention across both planned and short-notice sampling. All 18 participants completed the pre-scheduled baselines (At-Clinic and At-Home Pre-Event), while 16/18 completed the Post-Event Resampling despite receiving only one day’s notice of package delivery and a few days’ notice of sampling. The shelf stability of both Fellow and homeRNA kits allowed study packages to be prepared in advance and shipped as soon as sampling was initiated. Reliable overnight shipping was a critical component of the study protocol. This was especially important for the Fellow kits, which must arrive at the Fellow testing laboratory the day after collection to ensure successful processing.(25) While we also overnight shipped the blood samples, previous work has demonstrated that the stabilization method preserves RNA quality and yield for several days in transit, providing more flexibility.(28–31) Communication with participants also indicated that most were able to complete sample collection before work, suggesting that the protocol could be readily incorporated into typical weekday schedules.

In surveys, acceptability of protocols was scored similarly throughout the study, but there was a clear preference for sampling at home when it came to semen and blood collection. In survey responses regarding semen collection, participants were less likely to indicate that the test was awkward or embarrassing to perform when collecting at home. This aligns with previous literature which found that satisfaction levels were significantly higher for at-home collection of semen compared with in-clinic collection.(48) Further, participants rated understanding the instructions, completing collection, and returning the Fellow kit as easy, suggesting that mail in semen testing is most optimal for future studies.

Participants rated in-clinic and at-home use of the homeRNA platform similarly in terms of ease of use, discomfort, and collection time. Most participants reported no pain or only mild pain during blood collection, consistent with previous studies.(28–30,32,33) Although homeRNA has previously been paired with saliva collection in remote COVID-19 and long COVID studies,(33) this is the first study to combine remote blood sampling with at-home semen collection. This approach enables concurrent assessment of blood-based molecular biomarkers and measures of male reproductive health. Despite reporting similar experiences in both settings, participants preferred at-home sampling, primarily because of its convenience, and may support recruitment, retention, and protocol adherence in longitudinal studies requiring repeated biospecimen collection.

To our knowledge, this is the first study to integrate both PurpleAir and FAW devices in the same study. This advances our understanding of user experiences with PurpleAir and FAW, including acceptability in the context of clinical studies. Previous studies have incorporated PurpleAir monitors to characterize residential air quality; participant perspectives regarding the usability and acceptability of these devices have been well characterized.(38–41) In a longitudinal wildfire smoke study, Haack et al, collected homeRNA samples providing PurpleAir monitors to their participants, however usability and acceptability data was not collected.(32) The FAW has also been well characterized.(35,37,49) In this study, participants consistently rated the instructions for use and set-up of both platforms as very easy. However, for the PurpleAir monitors, participants were not uniformly in agreement that the monitor data was easy to understand, presenting an opportunity for future studies to include a data literacy component. Additionally, several participants disagreed or responded neutrally to the statements that the FAW was comfortable to wear and did not interfere with everyday activities. In the future, offering wristbands in multiple sizes may improve comfort. Further evaluation of participant experiences with PurpleAir and FAW could inform study protocols in future exposure studies.

This pilot study demonstrated several practical advantages that support the feasibility of a future remote longitudinal study. Targeted online ads reached men of reproductive age regardless of partnership status, sexual orientation, or reproductive intentions, broadening the pool of eligible participants and increasing the diversity of the cohort. Participants had the flexibility to sample and answer surveys when convenient without travel burdens. Providing the option to communicate with the study team via text message lowered communication barriers, enabling participants to ask questions and resolve concerns throughout the study. Passive environmental exposure monitoring also required minimal participant effort beyond setting up their PurpleAir Monitor and wearing their FAW (when applicable). Combined with minimally invasive at-home biofluid collection, may be benefits attributing to the low attrition rate.(50,51)

A major advantage of this study design is its ability to be deployed rapidly in response to transient environmental exposure events. Because study kits and collection reagents are shelf stable, they can be assembled and stored weeks or months in advance, minimizing the time between identifying an exposure event and shipping materials to participants. Leveraging existing overnight and expedited shipping networks enables participants in geographically dispersed locations to receive study kits within one to two days, allowing biospecimen collection while exposures are still occurring or shortly thereafter. This rapid deployment is critical for capturing acute biological responses, as time delays may miss short-lived environmental events and the molecular changes they induce. Although this protocol was developed with wildfire smoke exposure in mind, the same framework could be readily adapted to investigate other unexpected environmental or public health events, including chemical releases, industrial accidents, disasters, or other episodic exposures that require timely biospecimen collection.

Because this study was designed as a pilot to evaluate feasibility rather than estimate population-level effects, it included a relatively small cohort (target enrollment, N=20) with limited racial and ethnic diversity. These limitations will be addressed in future studies through expanded recruitment efforts and a larger, more diverse cohort. While the remote sampling protocol substantially reduced the need for in-person study visits, participants collecting blood, semen, and environmental exposure data were required to ship materials using two different shipping carriers. Future studies could further streamline participation by combining shipping methods across study materials or utilizing a centralized return process. Another challenge of the study design was prespecified wildfire smoke thresholds and limitations of current wildfire smoke forecasting. Study staff monitored ambient air quality in participants’ geographic regions and notified participants when a wildfire smoke event was anticipated. Because of the pilot nature of this study and timeframe for completion, a significant wildfire smoke event was not captured for the majority of participants.

In this feasibility and acceptability study, we demonstrated a novel remote protocol that integrates at-home semen and blood collection with residential and personal environmental exposure monitoring from baseline through periods of elevated exposure. Participants consistently preferred at-home rather than in-clinic sampling, highlighting the potential of remote study designs to reduce participant burden and support long-term retention. By demonstrating that repeated biospecimen collection and environmental exposure monitoring can be performed remotely with high participant acceptability, this study lays the groundwork for larger longitudinal investigations that would be difficult to conduct using traditional clinic-based approaches. In the future, a larger cohort spanning several regions that experience seasonal wildfire smoke exposure events that is completely remote would provide opportunity to improve our demographic diversity and increase the likelihood of capturing participant exposures thereby enabling robust investigation on how transient environmental exposures influence male reproductive health.

Future analyses from this cohort will integrate indoor air quality measurements from PurpleAir monitors, outdoor ambient air quality data, and personal exposure measurements from FAWs with semen quality metrics and blood transcriptomic profiles to investigate how short-term environmental exposures influence male reproductive health. Building on these findings, larger longitudinal studies could expand recruitment across multiple geographic regions that experience episodic declines in air quality, increasing the diversity of environmental exposures captured. The participant-friendly nature of this remote protocol, combined with its practicality for repeated biospecimen collection and rapid deployment, makes it well suited for longitudinal environmental health research. Because participants can self-collect and ship samples from virtually any location, this approach minimizes disruptions to daily life while improving the ability to capture time-sensitive exposure events. By enabling rapid, remote, and repeated collection of biospecimens alongside personal environmental exposure measurements, this platform provides a scalable framework for investigating the biological impacts of transient environmental exposures that are difficult to capture using conventional clinic-based study designs.

## CONCLUSION

We present a remote multi-biofluid and personal environmental monitoring protocol with the goal of assessing the effects of environmental exposures in reproductive aged men on semen quality. Using the Fellow Semen Analysis for semen collection, the homeRNAmax Blood Collection and Stabilization Kit for capillary blood sampling, and complimentary environmental monitoring with PurpleAir indoor PM_2.5_ monitors and Fresh Air Wristbands we evaluated the feasibility and acceptability of an entirely remote study protocol during a season with the potential for transient environmental exposure in the Pacific Northwest. Participants found biospecimen collection kits easy to use and preferred at-home versus travelling to a clinic supporting the practicality and participant-centered nature of this approach. Together, these findings demonstrate that integrating remote biospecimen collection with personal environmental exposure monitoring is a promising strategy for longitudinal reproductive health research.

## Supporting information

Supplemental Information for homeSPERM Feasibility and Acceptability Paper

## Data Availability

All data produced in the present study are available upon reasonable request to the authors.

## ACKNOWLEDGEMENTS

We want to thank our participants for volunteering their time and effort for this study, Connie Pete for initial sample processing and Kathy Winter for their assistance at the Center for Research in Reproduction and Contraception (CRRC) at the University of Washington.

This publication was supported by the National Institutes of Health (NIH) through the National Institute of General Medical Sciences award number R35GM128648 (for in-lab developments of homeRNA and Schmidt Sciences, LLC. Research was also supported by the University of Washington Pediatric and Reproductive Environmental Health Scholars K12 program (NIH K12ES033584, TMN trainee) and the National Center for Advancing Translational Sciences TL1TR002318 (ME, trainee). The REDCap used for human subjects consent and enrollment is supported by the Institute of Translational Health Sciences, which is funded by the National Center for Advancing Translational Sciences of the National Institutes of Health under award number UL1TR002319. Figure 1 was created in BioRender.com.

## NOTES

EB, AJH, and ABT filed patent 17/361,322 (Publication Number: US20210402406A1) and EB, AJH, FS, ME, and ABT filed patent 63/571,012 through the University of Washington on homeRNA and a related technology. AJH, LGB, and TMN report filing patents through the University of Washington. ABT reports filing multiple patents through the University of Washington and receiving a gift to support research outside the submitted work from Ionis Pharmaceuticals. EB has ownership in Salus Discovery, LLC, and Tasso, Inc. that develops blood collection systems used in this publication, and is employed by Tasso, Inc. Technologies from Salus Discovery, LLC are not included in this publication. EB is an inventor on multiple patents filed by Tasso, Inc., the University of Washington, and the University of Wisconsin-Madison. TMN has financial interests/equity in Tasso, Inc. and spouse is employed by Tasso, Inc. EB and ABT have ownership in Seabright, LLC, which will advance new tools for diagnostics and clinical research, including the homeRNA platform used in this publication, and EB and LGB are employed by Seabright, LLC. The terms of this arrangement for ABT and EB have been reviewed and approved by the University of Washington in accordance with its policies governing outside work and financial conflicts of interest in research. JFS is Chief Medical Officer at Fellow Health, Inc.

## Notes

### Author Declarations

This study was approved by the University of Washington Institutional Review Board (protocol 00022559).

## REFERENCES

1. Selevan SG, Borkovec L, Slott VL, Zudová Z, Rubes J, Evenson DP, et al. Semen quality and reproductive health of young Czech men exposed to seasonal air pollution. Environ Health Perspect 2000;108(9):887–94.

2. Hammoud A, Carrell DT, Gibson M, Sanderson M, Parker-Jones K, Peterson CM. Decreased sperm motility is associated with air pollution in Salt Lake City. Fertil Steril 2010;93(6):1875–9.

3. Zhou N, Cui Z, Yang S, Han X, Chen G, Zhou Z, et al. Air pollution and decreased semen quality: A comparative study of Chongqing urban and rural areas. Environ Pollut 2014;187:145–52.

4. Radwan M, Jurewicz J, Polańska K, Sobala W, Radwan P, Bochenek M, et al. Exposure to ambient air pollution-does it affect semen quality and the level of reproductive hormones? Ann Hum Biol 2016;43(1):50–6.

5. Jurewicz J, Radwan M, Sobala W, Brzeźnicki S, Ligocka D, Radwan P, et al. Association between a biomarker of exposure to polycyclic aromatic hydrocarbons and semen quality. Int J Occup Med Environ Health 2013;26(5):790–801.

6. Santi D, Vezzani S, Granata AR, Roli L, De Santis MC, Ongaro C, et al. Sperm quality and environment: A retrospective, cohort study in a Northern province of Italy. Environ Res 2016;150:144–53.

7. Xia Y, Han Y, Zhu P, Wang S, Gu A, Wang L, et al. Relation between Urinary Metabolites of Polycyclic Aromatic Hydrocarbons and Human Semen Quality. Environ Sci Technol 2009;43(12):4567–73.

8. Sokol RZ, Kraft P, Fowler IM, Mamet R, Kim E, Berhane KT. Exposure to Environmental Ozone Alters Semen Quality. Environ Health Perspect 2006;114(3):360–5.

9. Wu L, Jin L, Shi T, Zhang B, Zhou Y, Zhou T, et al. Association between ambient particulate matter exposure and semen quality in Wuhan, China. Environ Int 2017;98:219–28.

10. Omolaoye TS, Skosana BT, Ferguson LM, Ramsunder Y, Ayad BM, Du Plessis SS. Implications of Exposure to Air Pollution on Male Reproduction: The Role of Oxidative Stress. Antioxidants 2024;13(1):64.

11. Jurewicz J, Dziewirska E, Radwan M, Hanke W. Air pollution from natural and anthropic sources and male fertility. Reprod Biol Endocrinol 2018;16(1):109.

12. Ramsay JM, Fendereski K, Horns JJ, VanDerslice JA, Hanson HA, Emery BR, et al. Environmental exposure to industrial air pollution is associated with decreased male fertility. Fertil Steril 2023;120(3, Part 2):637–47.

13. Childs M, Martins M, Wilson A, Qiu M, Heft-Neal S, Burke M. Growing wildfire-derived PM2.5 across the contiguous U.S. and implications for air quality regulation. 2024 [cited 2026 Aug 20];Available from: https://eartharxiv.org/repository/view/8187/

14. Xie Y, Lin M, Horowitz LW. Summer PM2.5 Pollution Extremes Caused by Wildfires Over the Western United States During 2017–2018. Geophys Res Lett 2020;47(16):e2020GL089429.

15. Lindell LX, Holt SK, Petersen E, Gunaje N, Amighi A, Haack A, et al. Wildfire smoke exposure is associated with decreased sperm concentration and total motile sperm count. Fertil Steril 2026;125(2):316–25.

16. Ramsay JM, Horns JJ, Emery BR, Aston KI, Hotaling JM. EXPOSURE TO WILDFIRE SMOKE IS ASSOCIATED WITH DECREASED SEMEN QUALITY. Fertil Steril 2024;122(4):e210.

17. Rubin ES, Kornfield M, Parker P, Garg B, O’Leary T, Baldwin M, et al. Poor air quality from wildfire smoke is associated with decrease in total motile sperm count at time of intrauterine insemination. Fertil Steril 2024;121(5):881–3.

18. Godri Pollitt KJ. Beyond Inhalation: Air as an Integrative Matrix for Exposome Research. Environ Sci Technol 2026;60(38):26903–7.

19. Fyffe-Freil RC, Jannetto PJ, Vanderboom PM. To Boldly Go Where No Device Has Gone Before: Specimen Self-Collection for the Clinical Laboratory - ClinicalKey. Clin Microbilogy Newsl 2023;45(4):29–37.

20. Thangavelu MU, Wouters B, Kindt A, Reiss IKM, Hankemeier T. Blood microsampling technologies: Innovations and applications in 2022. Anal Sci Adv 2023;4(5–6):154–80.

21. Zarbl J, Eimer E, Gigg C, Bendzuck G, Korinth M, Elling-Audersch C, et al. Remote self-collection of capillary blood using upper arm devices for autoantibody analysis in patients with immune-mediated inflammatory rheumatic diseases. RMD Open [Internet] 2022 [cited 2026 June 1];8(2). Available from: https://rmdopen.bmj.com/content/8/2/e002641

22. Collier BB, Brandon WC, Chappell MR, Iacovetti G, Tokunaga N, Schaff UY, et al. Comparing capillary blood collection technologies: assessing patient experience, device performance, & clinical accuracy. Bioanalysis 17(21):1329–36.

23. Tolani AT, Morris JR, Nolte D, Kenfield SA, Belarmino A, Halpern JA, et al. HOW PATIENTS ARE UTILIZING MAIL-IN SEMEN ANALYSIS TESTING FOR FERTILITY EVALUATION; AN ASSESSMENT OF 3,159 PATIENTS SEEKING CARE ONLINE AND THROUGH UROLOGY, REI, OBGYN, AND WELLNESS PRACTICES. Fertil Steril 2024;122(4):e140–1.

24. Racicot K, Sakkas D, Barrett BC, Chiang K, Jenkins C. Validation of a mail-in delayed semen analysis protocol developed for home collection. FS Rep 2024;5(4):378–84.

25. Samplaski MK, Falk O, Honig S, Shin D, Matthews W, Smith JF. Development and validation of a novel mail-in semen analysis system and the correlation between one hour and delayed semen analysis testing. Fertil Steril 2021;115(4):922–9.

26. Wako WG, Clemens T, Ogletree S, Williams AJ, Jepson R. Validity, reliability and acceptability of wearable sensor devices to monitor personal exposure to air pollution and pollen: A systematic review of mobility based exposure studies. Build Environ 2025;277:112931.

27. Bernasconi S, Angelucci A, Aliverti A. A Scoping Review on Wearable Devices for Environmental Monitoring and Their Application for Health and Wellness. Sensors 2022;22(16):5994.

28. Eakman M, Stefanovic F, Berthier J, Robertson IH, Knudsen LA, Cardoso Carvalho C, et al. To the homeRNAmax: Developing an Improved Blood Self-Collection and Stabilization Platform for Remote Transcriptomic Studies. Anal Chem [Internet] 2026 [cited 2026 June 29];Available from: 10.1021/acs.analchem.5c07772

29. Haack AJ, Lim FY, Kennedy DS, Day JH, Adams KN, Lee JJ, et al. homeRNA: A Self-Sampling Kit for the Collection of Peripheral Blood and Stabilization of RNA. Anal Chem 2021;93(39):13196–203.

30. Brown LG, Haack AJ, Kennedy DS, Adams KN, Stolarczuk JE, Takezawa MG, et al. At-home blood collection and stabilization in high temperature climates using homeRNA. Front Digit Health [Internet] 2022 [cited 2025 July 16];4. Available from: https://www.frontiersin.org/journals/digital-health/articles/10.3389/fdgth.2022.903153/full

31. Stefanovic F, Brown LG, MacDonald J, Bammler T, Rinchai D, Nguyen S, et al. Your Blood is Out for Delivery: Considerations of Shipping Time and Temperature on Degradation of RNA from Stabilized Whole Blood. Anal Chem 2025;97(3):1635–44.

32. Haack AJ, Brown LG, Zeng Y, Khan T, Robertson IH, Kennedy DS, et al. A Flexible and Responsive Remote Study Design to Assess Gene Expression Changes During Wildfire Smoke Exposure with homeRNA, an At-home Blood Sampling Kit. MedRxiv Prepr Serv Health Sci 2025;2025.10.11.25337783.

33. Lim FY, Lea HG, Dostie AM, Kim S-Y, van Neel TL, Hassan GW, et al. homeRNA self-blood collection enables high-frequency temporal profiling of presymptomatic host immune kinetics to respiratory viral infection: a prospective cohort study. eBioMedicine 2025;112:105531.

34. Brown LG, Wei X, Milton LA, Alizai MY, MacDonald JW, Bammler TK, et al. From Home to Transcriptome: Comparing the Transcriptomic Profile of Induced Immune Response via Lipopolysaccharide Stimulation in homeRNA and Venous Blood. Anal Chem [Internet] 2026 [cited 2026 Jan 21];Available from: 10.1021/acs.analchem.5c06040

35. Lin EZ, Esenther S, Mascelloni M, Irfan F, Godri Pollitt KJ. The Fresh Air Wristband: A Wearable Air Pollutant Sampler. Environ Sci Technol Lett 2020;7(5):308–14.

36. Koelmel JP, Lin EZ, Nichols A, Guo P, Zhou Y, Godri Pollitt KJ. Head, Shoulders, Knees, and Toes: Placement of Wearable Passive Samplers Alters Exposure Profiles Observed. Environ Sci Technol 2021;55(6):3796–806.

37. Guo P, Lin EZ, Koelmel JP, Ding E, Gao Y, Deng F, et al. Exploring personal chemical exposures in China with wearable air pollutant monitors: A repeated-measure study in healthy older adults in Jinan, China. Environ Int 2021;156:106709.

38. Durkin A, Gonzalez R, Isaksen TB, Walker E, Errett NA. Establishing a Community Air Monitoring Network in a Wildfire Smoke-Prone Rural Community: The Motivations, Experiences, Challenges, and Ideas of Clean Air Methow’s Clean Air Ambassadors. Int J Environ Res Public Health 2020;17(22):8393.

39. Farooqui Z, Biswas J, Saha J. Long-Term Assessment of PurpleAir Low-Cost Sensor for PM2.5 in California, USA. Pollutants 2023;3(4):477–93.

40. Mullen CJ, Grineski SE, Herrera MM, Christian LP, Cuello J. “You Feel Empowered”: Parents’ Emotional Responses to Air Pollution Sensing at Home. J Health Soc Behav 2025;221465251406233.

41. Teigen KA, Cortes Espinosa M, McKinley H, Reed A, Berrang E, Ramos-Vazquez J, et al. A two-year indoor–outdoor monitoring study of household clean air interventions during ambient and wildfire smoke periods in King County, Washington. Environ Res Commun 2026;8(5):055027.

42. WHO laboratory manual for the examination and processing of human semen [Internet]. WHO Kobe Cent. [cited 2026 Mar 10];Available from: https://wkc.who.int/resources/publications/i/item/9789241547789

43. Olympic National Forest | Bear Gulch Fire | Forest Service [Internet]. fs.usda.gov. 2026 [cited 2026 Oct 27];Available from: https://www.fs.usda.gov/r06/olympic/fire/info/bear-gulch-fire

44. Sommer GJ, Wang TR, Epperson JG, Hatch EE, Wesselink AK, Rothman KJ, et al. At-home sperm testing for epidemiologic studies: Evaluation of the Trak male fertility testing system in an internet-based preconception cohort. Paediatr Perinat Epidemiol 2020;34(5):504–12.

45. Zhou J, Lin EZ, Wesselink AK, Schildroth S, Willis MD, Hatch EE, et al. Contribution of Consumer Products to Emerging Chemical Exposures in a Preconception Cohort Study. Environ Sci Technol 2026;60(29):20197–212.

46. Jensen TK, Jacobsen R, Christensen K, Nielsen NC, Bostofte E. Good Semen Quality and Life Expectancy: A Cohort Study of 43,277 Men. Am J Epidemiol 2009;170(5):559–65.

47. Chen T, Belladelli F, Giudice FD, Eisenberg ML. Male fertility as a marker for health. Reprod Biomed Online 2022;44(1):131–44.

48. Gao J, Duan Y-G, Yi X, Yeung WSB, Ng EHY. A randomised trial comparing conventional semen parameters, sperm DNA fragmentation levels and satisfaction levels between semen collection at home and at the clinic. Andrologia 2020;52(7):e13628.

49. Lin EZ, Herkert NJ, Hay D, Cooper EM, Stapleton HM, Godri Pollitt KJ. Comparative Assessment of Personal Exposures to Environmental Contaminants Using Silicone Wristbands and Fresh Air Wristbands. Environ Sci Technol 2026;60(37):26269–80.

50. Borg DJ, Haritopoulou-Sinanidou M, Gabrovska P, Tseng H-W, Honeyman D, Schweitzer D, et al. Barriers and facilitators for recruiting and retaining male participants into longitudinal health research: a systematic review. BMC Med Res Methodol 2024;24(1):46.

51. Gades NM, Jacobson DJ, McGree ME, Lieber MM, Roberts RO, Girman CJ, et al. Dropout in a longitudinal, cohort study of urologic disease in community men. BMC Med Res Methodol 2006;6(1):58.

