## Supplemental Information for homeSPERM Feasibility and Acceptability Paper for "Feasibility and Acceptability of a Prospective Remote Pilot Study for Assessment of Airborne Environmental Exposures and Male Fertility"

### "Male Remote Exposure Feasibility Study"


Ingrid H. Robertson[a*], Liddy Boland[a,b*], Damon Wing Hey Chan[a], Maximilian S. Jentzsch[c], Kathleen Maloney[d], Amanda J. Haack[a,e], Madeleine Eakman[a], Filip Stefanovic[a], Lauren G. Brown[a], Charles Tilander[d], Christine Loftus[d], Jane Edelson[f], Charles H. Muller[g], Erwin Berthier[a], Krystal J. Godri Pollitt[b], Nicole A. Errett[d], James F. Smith[h], Catherine J. Karr[d,f], John Amory[f], Ashleigh B. Theberge[a,c‡], Tristan Nicholson[c,d‡]

Affiliations

[a]Department of Chemistry, University of Washington, Seattle, Washington, USA

[b]Department of Environmental Health Sciences, Yale School of Public Health, New Haven, Connecticut, USA

[c]Department of Urology, School of Medicine, University of Washington, Seattle, Washington, USA

[d]Department of Environmental and Occupational Health Sciences, University of Washington, Seattle, Washington, USA

[e]Department of Chemical Engineering, Texas Tech University, Lubbock, TX, USA

[f]Department Medicine, University of Washington, Seattle, Washington, USA

[g]Department of Pediatrics, School of Medicine, University of Washington, Seattle, Washington, USA

[h]Department of Urology, University of California San Francisco, San Francisco, California, USA

[*]IR and LB contributed equally to this work



| | |
|---|---|
| SI.1 | The Fellow Semen Analysis Kit (Fellow) |
| SI.2 | homeRNAmax Blood Collection and Stabilization Kit (homeRNA) |
| SI.3 | Fresh Air Wristband (FAW) |
| SI.4 | PurpleAir Touch |
| SI.5 | Compounds found on the Fresh Air Wristbands |
| SI.6 | Exposure Table |
| Si.7 | Lifestyle Table |

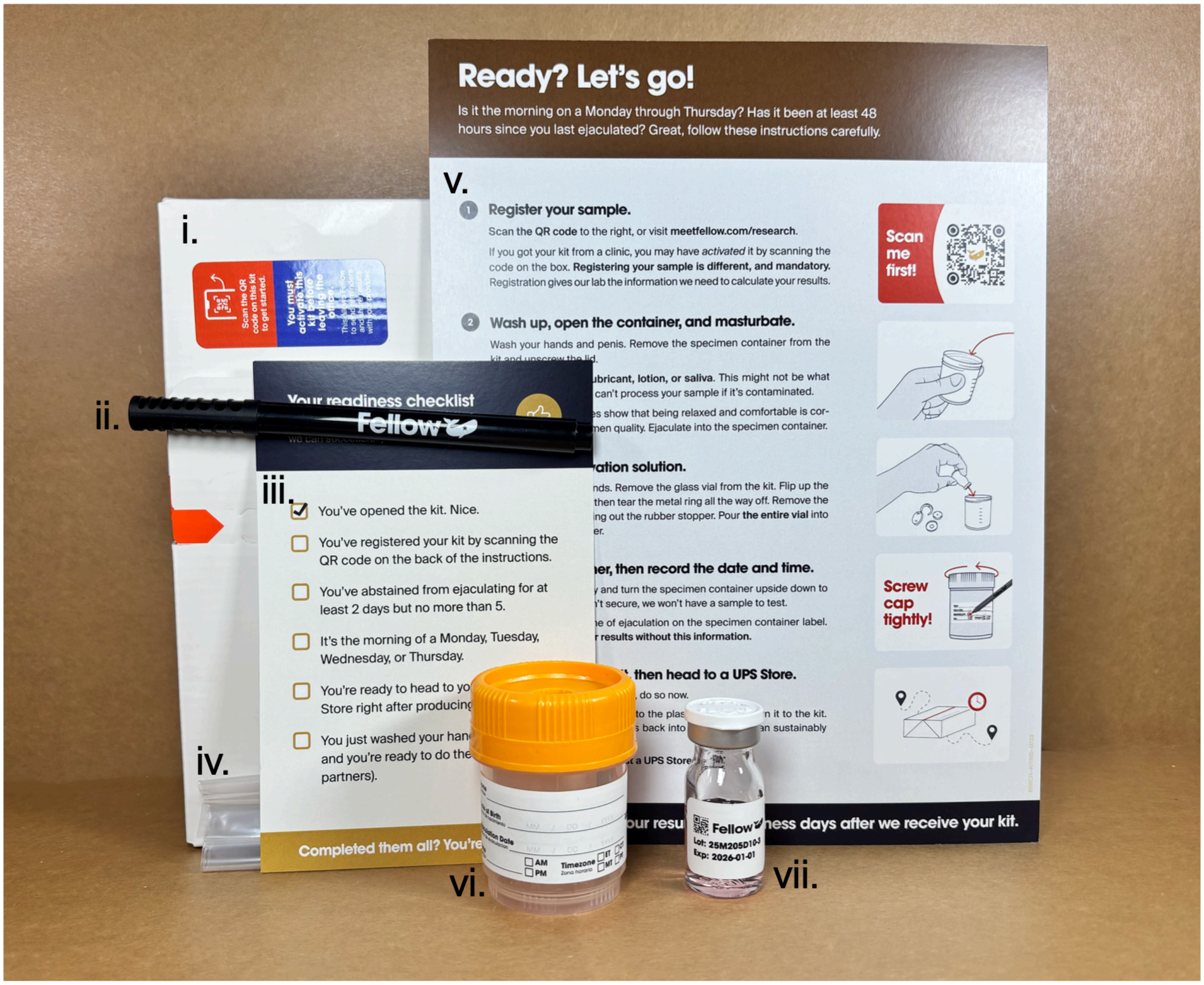


**SI.1 The Fellow Semen Analysis Kit (Fellow)** is a direct-to-consumer platform for at-home semen collection and mail-in laboratory analysis. Participants abstain from ejaculation for 2-5 days, collect a semen sample directly into the provided specimen cup, add a stabilizing reagent, and ship the sample overnight to the Fellow laboratory. After the lab performs a semen analysis, resulting data is run through a proprietary predictive model that extrapolates semen parameters at the time of the ejaculation. The kit includes (i.) a pre-labeled box to ship the sample, (ii.) a pen, (iii.) a sampling readiness checklist, (iv.) a plastic bag to hold the specimen cup after collection, (v.) instructions for use, (vi.) a specimen cup, and (vii.) stabilization fluid. (1)

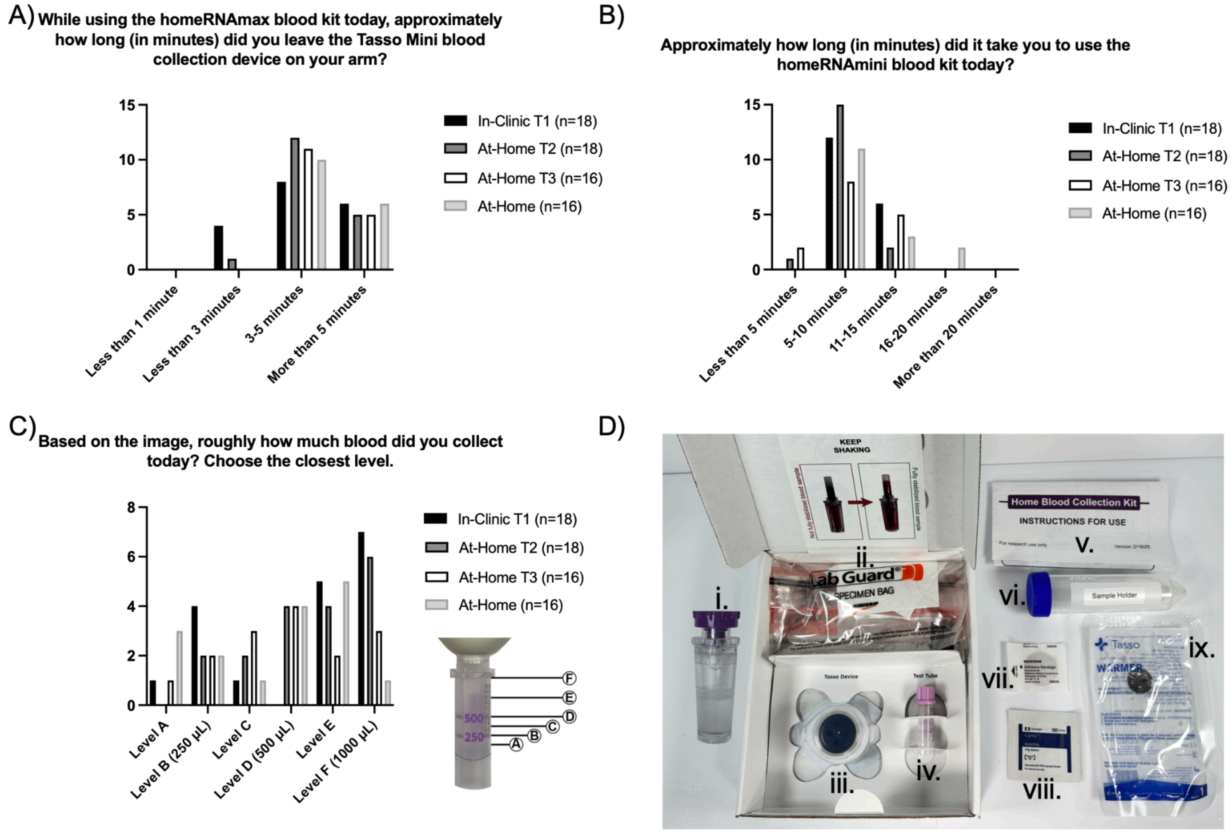


**SI.2 homeRNAmax Blood Collection and Stabilization Kit (homeRNA):** The homeRNA kit is designed to stabilize RNA at ambient temperatures for shipping, sometimes taking multiple days, maintaining RNA integrity for downstream transcriptomic analysis. (A) Length of time participants collected blood using the Tasso Mini, instructions indicated 5 minutes or until blood reached the collar or "F" on the BD microtainer (1 mL). (B) Length of time it took to use the entire kit, which included heating the arm, sanitizing the area, blood collection, and stabilization. (C) Visual approximation by participants of total blood volume collected before stabilization. (D) The homeRNAmax Blood Collection and Stabilization Kit (homeRNA) allows for RNA stabilization of up to 1 mL of capillary blood collected from the upper arm. . From left to right included in the homeRNA kit: (i.) homeRNA stabilizer tube, (ii.) specimen bag, (iii.) Tasso Mini, (iv.) EDTA coated blood tube (BD Microtainer), (v.) instructions for use, (vi.) 50-mL conical tube, (vii.) circular bandage, (viii.) alcohol swab, and (ix.) Tasso branded warmer. (2–8)

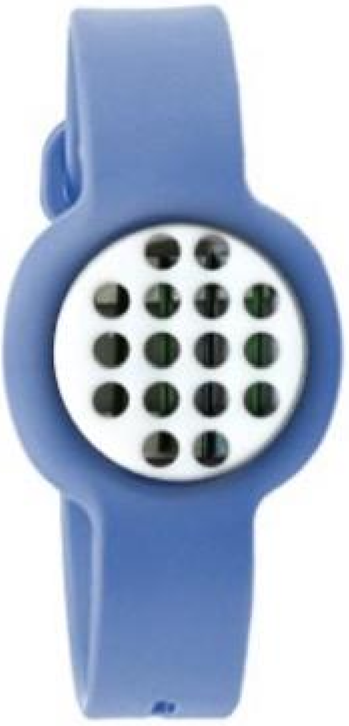

**SI.3 Fresh Air Wristband (FAW):** Participants wore the Fresh Air Wristbands continuously for seven days leading up to sample collection. Fresh Air wristbands are low-cost, wearable passive samplers that collect airborne contaminants onto custom fabricated polymeric membrane sorbent bars comprised of polydimethylsiloxane (PDMS).(9) Chemical analysis of the PDMS sorbent bars via by thermal desorption gas chromatography high-resolution mass spectrometry provides a time-integrated exposure measurement.(9,10) To quantify exposure to airborne compounds, Fresh Air wristbands were utilized to evaluate exposures to several chemical groups including polycyclic aromatic hydrocarbons (PAHs), phthalates, pesticides, and flame retardants.(11)

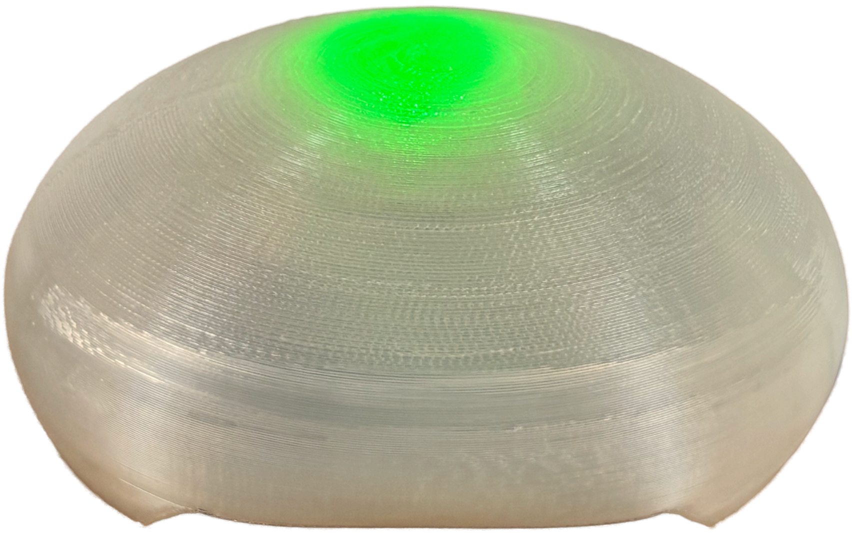

**SI.4 PurpleAir Touch**: The PurpleAir Monitor is a low-cost, commercially available air-quality monitor that has been deployed globally since 2017. This study used the PurpleAir Indoor Air Quality Monitor to obtain individual indoor air quality measurements. For outdoor air quality, existing PurpleAir sensors were used to estimate outdoor air quality for each participant covering the period from 72 days prior to enrollment through the end of the study to capture the full spermatogenesis cycle.(12)

**SI. 5 Exposures found on the Fresh Air Wristbands:** Nine participants (50%) were selected to wear a Fresh Air Wristband. All nine completed baseline sampling, and six completed repeat sampling, yielding 15 wristband samples. Thirty-nine compounds were detected across the wristbands: 10 phthalates, four polycyclic aromatic hydrocarbons, five flame retardants, eight pesticides, two UV filters, two musks, one nitro compound, five uncategorized compounds, THC, and nicotine.

| EXPOSURES OVER STUDY PERIOD | N=18(%) |
|---|---|
| Does your occupation require you to work outside? | |
| Yes | 2(11) |
| No | 13(72) |
| Sometimes | 3(17) |
| In the past three months, have you used or experienced any of the following?* | |
| Cigarettes, Joints, Blunts | 3(17) |
| Marijuana or Cannabis | 7(39) |
| Secondhand Smoke Exposure | 5(28) |
| In the past week, did you engage in exercise outdoors?* | |
| Yes | 16(89) |
| No | 2(11) |
| Estimate the amount of outdoor exercise time in the last two weeks | |
| Less than 10 minutes | 0 |
| 10 min – 1 hour | 6(33) |
| 1-2 hours | 4(22) |
| 2-3 hours | 4(22) |
| More than 3 hours | 2(11) |
| In the past three months, estimate the amount of time that you spent outside, on average, on a typical day* | |
| Less than 10 minutes | 0 |
| 10 min – 1 hour | 3(17) |
| 1-2 hours | 7(39) |
| 2-3 hours | 2(11) |
| More than 3 hours | 6(33) |
| In the past three months, did you experience wildfire smoke exposure? | |
| Yes | 0 |
| No | 14(78) |
| Unsure | 4(22) |
| During the duration of the study, did you attend any of the following events or use the following equipment? | |
| Gas Stove, Indoors | 7(39) |
| Barbecue Grill (Propane, Charcoal, Other) | 4(22) |
| Bonfire/Campfire | 7(39) |
| Fireworks Show | 2(11) |
| Do you use ventilation with the stove? | |
| Yes | 7(100) |
| No | 0 |

**SI. 6 Exposures Table:** Self-reported exposure/lifestyles of participants during the study period, asked at the end of the Timepoint 4.

| LIFESTYLE AT SAMPLE TIMEPOINTS | N = 18 (%) Clinic Timepoint 1 | N=18 (%) At-Home Timepoint 2 | N = 16 (%) At-Home Timepoint 3 | N=16 (%) At-Home Timepoint 4 |
|---|---|---|---|---|
| In the past three months, have you used or experienced the following? | | | | |
| Cigarettes, Joints, Blunts | (17) | 4(22) | | |
| Marijuana or Cannabis | (39) | 4(22) | | |
| Secondhand Smoke Exposure | (28) | 6(33) | | |
| In the past week, did you engage in exercise outdoors? | | | | |
| Yes | (89) | 14(78) | 7(44) | 9(50) |
| No | (11) | 4(22) | 9(56) | 7(39) |
| Estimate the amount of outdoor exercise time in the last 2 weeks | | | | |
| Less than 10 minutes | | 0 | 0 | 0 |
| 10 min – 1 hour | (38) | 5(36) | 4(57) | 2(22) |
| 1-2 hours | (25) | 5(36) | 0 | 2(22) |
| 2-3 hours | (25) | 2(14) | 0 | 2(22) |
| More than 3 hours | (13) | 2(14) | 3(43) | 3(33) |
| In the past three months, did you experience wildfire smoke exposure? | | | | |
| Yes | | 3(17) | 9(56) | 8(44) |
| No | (78) | 11(61) | 1(6) | 4(22) |
| Unsure | (22) | 4(22) | 6(38) | 4(22) |
| In the past three months, did you take any of the following protective measures to lessen your exposure to wildfire smoke or other air quality issues? | | | | |
| Use of an air purifier at home or work | 4(22) | 4(22) | 2(13) | 3(19) |
| MERV13 or higher filter on HVAC system at home or work | 2(11) | 1(5) | 1(6) | 1(6) |
| Closing windows and doors | 5(28) | 1(5) | 7(44) | 9(56) |
| Limiting time spent outdoors | 3(17) | 3(17) | 5(31) | 5(31) |
| Limiting time spent outside exercising | 3(17) | 2(11) | 4(25) | 3(19) |
| Use of N95 mask when outside | 2(11) | 0 | 1(6) | 1(6) |
| Other | 1(6) | 0 | 0 | 0 |
| None | 10(56) | 10(56) | 7(44) | 7(44) |

**SI. 7 Lifestyle Table:** These questions were asked in a survey at each sample timepoint.
